# Mathematical analysis of Córdoba calcifediol trial suggests strong role for Vitamin D in reducing ICU admissions of hospitalized COVID-19 patients

**DOI:** 10.1101/2020.11.08.20222638

**Authors:** Irwin Jungreis, Manolis Kellis

## Abstract

A randomized controlled trial of calcifediol (25-hydroxyvitamin D_3_) as a treatment for hospitalized COVID-19 patients in Córdoba, Spain, found that the treatment was associated with reduced ICU admissions with very large effect size and high statistical significance, but the study has had limited impact because it had only 76 patients and imperfect blinding, and did not measure vitamin D levels pre- and post-treatment or adjust for several comorbidities. Here we reanalyze the reported results of the study using rigorous and well established statistical techniques, and find that the randomization, large effect size, and high statistical significance address many of these concerns. We show that random assignment of patients to treatment and control groups is highly unlikely to distribute comorbidities or other prognostic indicators sufficiently unevenly to account for the large effect size. We also show that imperfect blinding would need to have had an implausibly large effect to account for the reported results. Finally, comparison with two additional randomized clinical trials of vitamin D supplementation for COVID-19 in India and Brazil indicates that early intervention and rapid absorption may be crucial for the observed benefits of vitamin D. We conclude that the Córdoba study provides sufficient evidence to warrant immediate, well-designed pivotal clinical trials of early calcifediol administration in a broader cohort of inpatients and outpatients with COVID-19.

## Introduction

As of this writing, the 2019-2020 COVID-19 pandemic has resulted in more than one million deaths worldwide, and at times overloaded hospital and intensive care unit (ICU) capacities in the worst-affected areas. Effective treatments to decrease the severity of the disease are urgently needed.

Several lines of evidence have suggested a relationship between the vitamin D endocrine system and both the incidence and severity of COVID-19 [1]. Here we focus on the latter. Most of the risk factors known to be associated with poor COVID-19 outcome [2,3] are also risk factors for vitamin D (25-hydroxyvitamin D (25OHD)) deficiency [4,5], including advanced age, obesity, darker skin, hypertension, cardiovascular disease, kidney disease, and diabetes. European countries with higher 25OHD deficiency tended to have higher COVID-19 mortality rates [6,7], and an analysis of 117 countries found that those with high northern latitude, which presumably correlates with lower UVB light and lower 25OHD serum levels, have higher mortality rates when adjusted for age [8].

Serum 25OHD levels were predictive of COVID-19 severity and mortality in several cohort and retrospective studies. A retrospective study of 80 patients with confirmed COVID-19 in Madrid, Spain, found that vitamin D deficiency measured within the three preceding months predicted more severe disease [OR 3.2, (95% CI: 0.9-11.4), p = 0.07] [9]. A cohort study found that 17 hospitalized patients ≥50 years of age with COVID-19 admitted to a Singapore hospital after a treatment regimen with vitamin D, magnesium, and vitamin B_12_ was initiated had significantly less need for oxygen or ICU admission than the 26 patients admitted before the regimen was initiated (17.6% versus 61.5%, p = 0.006), and the difference remained substantial and significant when adjusted for age or hypertension [10]. A study in Heidelberg, Germany, of 185 COVID-19 patients found 25OHD levels below 12 ng/mL were significantly associated with higher risk of the need for mechanical ventilation (HR 6.12, 95% CI 2.79–13.42, p < 0.001) or death (HR 14.73, 95% CI 4.16–52.19, p < 0.001, respectively) when adjusted for age, gender, and comorbidities [11]. A retrospective observational study of 149 COVID-19 patients in Istanbul, Turkey found that 25OHD serum levels were lower in patients with severe illness, and found that vitamin D levels independently predicted mortality after adjusting for age and several comorbidities [12]. A retrospective study in a Mexican hospital found that among 172 hospitalized COVID-19 patients, those with 25OHD serum levels below 8 ng/mL had 3.68 higher risk of dying from COVID-19 than those with higher levels, though the study did not control for possible confounders [13].

Several mechanisms have been proposed for how activation of the vitamin D receptor (VDR) could decrease acute lung injury and Acute Respiratory Distress Syndrome (ARDS), which are the major factors determining poor prognosis of hospitalized COVID-19 patients [14–16], namely decreasing cytokine storm, modulating neutrophil activity, maintaining the pulmonary epithelial barrier, stimulating epithelial repair, and decreasing hypercoagulability and thrombosis [17,18]. An analysis of gene expression differences between COVID-19 patients and controls found that dysregulation of the Renin Angiotensin System (RAS) in COVID-19 patients leads to increased levels of bradykinin (a “bradykinin storm”), which could account for many symptoms of ARDS, suggesting this could be corrected by vitamin D supplementation to decrease levels of renin [19].

High dose vitamin D_3_ supplementation has been found to be safe in critically ill patients with low 25OHD serum levels. The consensus recommendation of the European Society for Clinical Nutrition and Metabolism is that a high dose of vitamin D_3_ (500,000 IU or, equivalently, 12,500 mcg) can be administered to critically ill patients with low plasma vitamin D levels, based on seven randomized trials of 716 critically ill adult patients that found no side effects in six months of follow up after supplementation with 200,000-540,000 IU (5,000-13,500 mcg) [20].

A small randomized controlled trial in Córdoba, Spain, of calcifediol (25-hydroxyvitamin D_3_, or 25(OH)D_3_) for hospitalized COVID-19 patients (henceforth, “the Córdoba study”) found dramatic reduction in the need for ICU admission [21]. This study has been viewed as a small preliminary study, suggesting at most that further study might be warranted. It has gotten relatively little attention, though its strengths and weaknesses were discussed in this article [22], and a Bayesian cost/benefit analysis found the expected benefits, in terms of lives saved and severe illness avoided, of immediately adopting the treatment protocol were considerably higher than the expected costs [23]. Two other randomized trials of vitamin D formulations for COVID-19 had mixed results: a small trial in India found a high dose of vitamin D_3_ shortened time to viral clearance in asymptomatic or mildly symptomatic SARS-CoV-2 individuals [24], and a larger study in Sao Paulo, Brazil did not find a statistically significant benefit of a high dose of vitamin D_3_ in hospitalized COVID-19 patients [25].

Here we apply rigorous and well established statistical techniques to reanalyze the results of the Córdoba study. We show that the randomization and large effect size address many of the concerns that have been raised about the study, including the small study size, the possibility that the results are due to uneven distribution of comorbidities between treated and control groups, and imperfect blinding. Specifically, we show that the probability of obtaining such a large effect size by chance if the treatment had no effect is less than one in a million, that the probability that randomization would produce a difference in comorbidities between the treatment and control groups large enough to account for the effect is less than one in 60,000, and for the results to be due to imperfect blinding would have required placebo and related effects to cause an implausibly large risk reduction. The differences in results between the India and Córdoba studies vs. the Sao Paulo study indicate that early application of the treatment is critical. We investigate the question of whether the Córdoba results will generalize to different cohorts, and consider next steps.

## Results and Discussion

### The Córdoba calcifediol study

The Córdoba study was a randomized, controlled, partly-masked clinical trial at Reina Sofia University Hospital in Córdoba, Spain, to test whether treatment with calcifediol could decrease the need for ICU admission among hospitalized COVID-19 patients, which would also likely decrease the risk of death. The study was pre registered (NCT04366908), with ICU admission and death as the primary outcome measures. The study report was ambiguous with regard to who was masked; we discuss this in the section on blinding.

Electronic randomization by hospital statisticians assigned 76 consecutive hospitalized confirmed COVID-19 patients to treatment or control groups in a two-to-one ratio. All received the hospital standard of care at the time, which was hydroxychloroquine and azithromycin. Treated patients also received oral calcifediol consisting of one 532 mcg soft capsule on the day of admission and 266 mcg on days 3 and 7, then weekly until discharge. Calcifediol was used rather than alternative vitamin D formulations because of its reliable intestinal absorption and rapid restoration of serum concentration. The dosage is well within the safe dosage guidelines for critically ill patients with low 250HD plasma levels (12,500 mcg as a single dose) from the European Society for Clinical Nutrition and Metabolism [20], even when adjusted for the higher absorption of calcifediol compared to vitamin D_3_. ICU admission was determined by a blinded multidisciplinary selection committee based on previously specified criteria.

A dramatic decrease in the need for ICU admission was observed in the treatment group (1 out of 50, 2%) as compared to the control group (13 out of 26, 50%). In order to determine if this difference was due to different characteristics of the patients in the two groups, the authors reported statistics for 15 prognostic risk factors (**Table 1**), and used a multivariate logistic regression to compute the adjusted odds ratio correcting for the two risk factors, hypertension and type 2 diabetes mellitus, that were significantly higher in the control group. After correcting for these imbalances, ICU admissions were still dramatically lower among the treated patients (odds ratio 0.03, 95% CI: 0.003-0.25).

**Table 1.** Prognostic factors for COVID-19 at baseline. Entries in black are taken from Castillo et al Table 2 [21], reproduced here for convenience. Entries in blue are comparison of the overall Córdoba cohort to hospitalized COVID-19 patients in the SOLIDARITY study [37], including the number and percent of patients having each characteristic measured in both studies for the overall cohorts, expected number in the Córdoba study based on percentage in the SOLIDARITY study, and p-value for the difference. Numbers for “Male” are from Castillo et al Table 1. For age intervals matching those reported in the SOLIDARITY study (marked by *), the Córdoba counts were estimated using normal distributions with means and standard deviations matching those reported in the Córdoba study. One additional significant digit of the p-value for high blood pressure has been included because we make use of it in the text.

| Poor prognosis risk factor | Group receiving Calcifediol (n=50) | Group without Calcifediol (n=26) | IC 95% | P | SOLIDARITY (n=11266) | Córdoba Total (n=76) | Córdoba expected | Cór vs SOL P-val |
| --- | --- | --- | --- | --- | --- | --- | --- | --- |
| ≥ 60 years | 14 (28%) | 5 (19.23%) | -0.11 – 0.28 | 0.40 |  |  |  |  |
| Previous lung disease | 4 (8%) | 2 (7.69%) | -0.12 – 0.13 | 0.96 | 635(5.64%) | 6(7.89%) | 4 | 0.257 |
| Previous Chronic kidney disease | 0 | 0 | - | - |  |  |  |  |
| Previous Diabetes mellitus | 3 (6%) | 5 (19.23%) | -0.30 – 0.03 | 0.08 | 2768(24.57%) | 8(10.53%) | 19 | 0.002 |
| Previous High blood pressure | 11 (24.19%) | 15 (57.69%) | -0.58 – -0.13 | 0.0023 |  |  |  |  |
| Previous Cardiovascular disease | 2 (4%) | 1 (3.85%) | -0.09 – 0.09 | 0.97 | 2337(20.74%) | 3(3.95%) | 16 | <0.001 |
| Immunosuppressed & transplanted | 6 (12%) | 1 (3.85%) | -0.03 – 0.20 | 0.24 |  |  |  |  |
| At least one prognostic bad risk factor* | 24 (48%) | 16 (61.54%) | -0.37 – 0.10 | 0.26 |  |  |  |  |
| PaO2/FiO2 (mean +/- SD) | 346.57 +/- 73.38 | 334.62 +/- 66.33 | -22.29 – 46.19 | 0.49 |  |  |  |  |
| C-reactive protein (mg/L) (mean +/-SD) | 82.93 +/- 62.74 | 94.71 +/- 63.64 | -42.15 – 18.59 | 0.44 |  |  |  |  |
| LDH (U/L)(mean +/-SD) | 308.12 +/- 83.83 | 345.81 +/- 108.57 | -82.46 – 7.08 | 0.10 |  |  |  |  |
| D-Dimer (ng/mL) (mean +/-SD) | 650.92 +/- 405.61 | 1333.54 +/- 2570.50 | -360.29 – 1725.53 | 0.19 |  |  |  |  |
| Lymphocytes < 800/μL | 10 (20%) | 6 (23.08%) | -0.16 – 0.23 | 0.75 |  |  |  |  |
| Ferritin (ng/mL) (mean +/-SD) | 691.04 +/- 603.54 | 825.16 +/- 613.95 | -166.31 – 434.55 | 0.36 |  |  |  |  |
| IL-6 (22/48) (pg/mL) (mean +/-SD) | 28.88 +/- 75.05 | 19.54 +/- 19.45 | -41.88 – 23.19 | 0.41 |  |  |  |  |
| Male | 27(54%) | 18(69.23%) |  | 0.15 | 6985(62.00%) | 45(59.21%) | 47 | 0.348 |
| <50 years* | 18(36%) | 9(34.62%) |  | 0.56 | 3995(35.46%) | 27(35.53%) | 27 | 0.538 |
| 50-69* | 28(56%) | 16(61.54%) |  | 0.41 | 5125(45.49%) | 44(57.89%) | 35 | 0.020 |
| 70+* | 3(6%) | 1(3.85%) |  | 0.58 | 2146(19.05%) | 4(5.26%) | 14 | 0.001 |

The mortality rate among treated patients was also lower (0 out of 50 treated patients, 0%, versus 2 out of 26 control patients, 8%). The number of deaths was too small to achieve statistical significance against a null hypothesis of no effect (one-sided hypergeometric p = 0.11) but the result is consistent with the plausible hypothesis that the decrease in mortality would be similar to the decrease in ICU admissions.

### Statistical significance is extremely high

Although the study included a small number of patients in absolute terms, the very large effect size observed allows us to make confident inferences.

The primary concern with a small trial is that differences in the outcomes of the treated and control groups could be due not to the treatment but instead due to chance, under the null hypothesis that assignment to the treatment group has no effect. However, the number of patients needed to rule out this null hypothesis depends on the size of the effect, with large studies being necessary for subtle effects but a smaller number of patients being sufficient to detect a more robust effect. The probability that an effect as large as the one observed could arise due to chance under the null hypothesis, the p-value, tells us whether the number of patients in the study provides enough statistical power for the observed effect size.

The authors of the study calculated the statistical significance of the lower rate of ICU admission in the treatment group as compared to the control group using the *χ*^2^ approximation, and reported a p-value p<0.001, but we find that the exact p-value is much smaller, indicating much greater statistical significance. Since 14 of the 76 patients in the study required ICU admission, and under the null hypothesis the probability of a patient requiring ICU admission does not depend on whether the patient is in the treated or control groups, the one-sided p-value is, by definition, the probability that if we randomly choose 14 out of the 76 patients, one or fewer of them would be in the treatment group. That probability is given by the hypergeometric distribution, the one-sided version of Fisher’s exact test, which is defined as the probability that among *n* elements randomly drawn from a finite population without replacement, *k* of them will have a specified feature. In our case, the random draws are the 14 out of 76 patients requiring ICU admission, and the specified feature is being in the treatment group. Using the hypergeometric distribution, we find that the p-value is p = 0.00000077 = 7.7 x 10^−7^. A one-sided p-value is appropriate, because the hypothesis being tested is that calcifediol *decreases* disease severity, rather than that it has some effect in either direction.

There is a qualitative difference between results having a p-value of 0.001 and a p-value less than 10^−6^. As of April 30, 2020 when the Córdoba study was registered with clinicaltrials.gov, there were approximately 500 randomized intervention studies of COVID-19 treatments registered there. Even if none of those treatments were effective, one would expect approximately one of them to obtain a p-value as low as 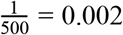 simply due to chance, and it is plausible that one of them would obtain a p-value less than 0.001. P-hacking and HARKing (hypothesizing after results are known) effectively allow several hypotheses to be tested at once, with only the most significant result being reported [26]. However, these effects are severely limited by preregistration, since the hypothesis being tested is specified before the trial is conducted. It is not plausible that in a preregistered clinical trial with a single intervention, a combination of these effects could explain a p-value less than 10^−6^, which is less than one thousandth of the smallest p-value that would be expected by chance among 500 studies if the intervention had no effect. In short, we can be confident that if assignment to the treatment group had no effect, we would not have observed these results simply due to chance.

### Randomization protects against comorbidities and other prognostic risk factors

A related concern is that the control group could have been enriched for comorbidities or other prognostic risk factors that make a severe outcome more likely. As noted above, the authors found that the effect size and statistical significance remained high (odds ratio 0.03, 95% CI: 0.003-0.25) after correcting for the two prognostic risk factors that were significantly enriched in the control group, namely hypertension and type 2 diabetes mellitus, out of the fifteen risk factors that were measured (**Table 1**), but other prognostic risk factors such as obesity and overall disease severity at hospital admission were not reported. It is generally recommended that all known confounders be checked and their effects modeled, but the concerns that such explicit modeling are meant to address are alleviated for the Córdoba study by the large effect size and statistical significance.

In a retrospective study, it is essential to ensure that the treated and control groups are well matched for known prognostic risk factors, and confounding by *unknown* risk factors can invalidate the results. However, in a randomized study with large effect size and statistical significance, the randomization protects against that. On the one hand, a very large imbalance in prognostic risk factors would be needed to explain the factor of 25 difference in relative risk of ICU admission observed in the Córdoba study. This follows from Cornfield’s Inequalities [27], from the seminal paper that established the causal link between smoking and lung cancer [28]. Intuitively, a difference of, say, three-fold in the rates of some risk factor such as obesity between the treatment and control groups cannot explain more than a three-fold difference in outcomes. That is because if a risk factor were 100% causal and there were no other factors affecting the outcome then a three-fold difference in the rates of that risk factor would explain exactly a three-fold difference in the outcomes, and the effect of that risk factor on outcomes will be diluted if it is less than 100% causal and there are other factors affecting the outcome. On the other hand, randomization limits how large an imbalance is likely to occur: for example, if we flip a coin 50 times we will get 5 or fewer heads only about twice in a billion attempts (p = 2.1 x 10^−9^).

There can be any number of prognostic risk factors, but if we knew what all of them were, and their effect sizes, and the interactions among them, we could combine their effects into a single number for each patient, which is the probability, based on all known and yet-to-be discovered risk factors at the time of hospital admission, that the patient will require ICU care if not given the calcifediol treatment. Call this (unknown) probability P_prognostic_(Patient). Even though we do not know the actual values of these probabilities, we will show that we can make some inferences about them. While knowing the particular comorbidities that contribute can be valuable for understanding the mechanism of the disease, only the combined probability is needed for determining whether they explain the difference in outcomes between the treated and control groups. It might seem counterintuitive that we can use a single number to replace a large number of prognostic risk factors, some of which could have large imbalances. For example, some individual risk factor such as obesity could be substantially overrepresented in the control group. However, looking at one risk factor at a time does not give the complete picture. A large overrepresentation of obesity in the control group might be counterbalanced by a large overrepresentation of elderly patients in the treatment group, or by the combined effect of several smaller overrepresentations in the treatment group. It is only by combining the effects of all risk factors that we can accurately judge whether one group of patients or the other has a greater predisposition to severe disease.

We can infer something about how the values of P_prognostic_(Patient) are distributed between the treatment and control groups from the fact that patients were assigned to these groups randomly. If we knew this distribution, it would be possible to calculate how likely it would be that we would observe a difference in ICU admissions as large as the actual observed difference, under the null hypothesis that assignment to the treatment group has no effect. If this likelihood were larger than our significance threshold, say 0.05, then that would mean that the result would no longer be statistically significant after adjusting for all prognostic risk factors. The following mathematical theorem allows us to estimate how likely it is that the randomization would distribute the prognostic risk factors so unevenly between the treatment and control groups that after adjusting for them the result would no longer be statistically significant.

#### Theorem

In a randomized controlled study, let *p* be the p-value of the study results, and let *q* be the probability that randomization would distribute prognostic risk factors sufficiently unevenly between the treatment and control groups that after controlling for them the results of the study would no longer be statistically significant at the 95% level. Then 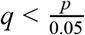.

We supply the proof in Methods. The basic idea is that if the difference in outcomes results from a chance imbalance of prognostic risk factors, that is a special case of the difference in outcomes resulting from chance. The only assumption is that the randomization is truly random, i.e., that all patients have the same probability of being assigned to the control group, and that these assignments are independent for different patients. It does not rely on any assumption about the number or effect sizes of the prognostic risk factors. Applying this theorem, we find that 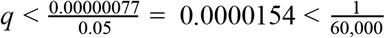.

So, there is less than a one in 60,000 chance that the randomization would have produced a distribution of comorbidities or other prognostic risk factors so uneven that the results would no longer have been significant at the 95% level. Similarly, there is less than a one in 12,000 chance of that at the 99% significance level. To determine the probability that the randomization actually did produce such an uneven distribution of prognostic risk factors, it is appropriate to condition on the actual values of any risk factors that were measured, namely the ones in **Table 1**. Castillo et al provided only summary statistics, which is insufficient information to calculate the conditional probability exactly, but we gain some assurance because: first, adjusting for the two risk factors that were significantly enriched in the control group (hypertension and diabetes), did not dramatically change the odds ratio (0.03 instead of 0.02); second, age, which is known to be the most important risk factor for severe COVID-19, is enriched in the treatment group; and finally, other prognostic risk factors are not very different between the two groups or are enriched in the treatment group.

The presence of comorbidities was among the criteria used for ICU admission, so one might wonder whether an imbalance in comorbidities could have led to increased ICU admission for this reason irrespective of their effect on disease severity. However, whether comorbidities led directly to increased ICU admission via the admission criteria or indirectly through disease severity, the math is the same, so the above analysis addresses this case as well.

In summary, the randomization and high statistical significance allow us to confidently rule out imbalances in comorbidities or other prognostic risk factors, whether measured or not, as the explanation for the large difference in ICU admissions between the treated and control groups that was observed in the study.

### Large effect size protects against imperfect blinding

We have defined the null hypothesis to be that assignment to the treatment group had no effect, but the hypothesis we really want to rule out is that the *treatment itself* had no effect. These are different because assignment to the treatment group could have effects other than the treatment itself, such as placebo effect. The purpose of blinding and placebo control is to eliminate any such effects. The Córdoba study was performed during the peak of COVID-19 infections in Spain which overwhelmed many health care facilities, so a full double blinded placebo-controlled trial with extensive blood sampling was not feasible. However, steps were taken to minimize non-treatment effects of being in the treatment group. We describe these below, based on our conversations with Dr José Manuel Quesada Gómez, senior author of the Córdoba study.

Most importantly, the decision of whether a patient should be admitted to the ICU was made by a committee of specialists who *were* blinded, and that decision was based on the prespecified hospital protocol. The data collectors and statisticians were also blinded, eliminating those potential sources of bias. Although no placebo was used, patients were not told which group they were in, and because they were receiving several other drugs as part of the hospital standard of care, and were hospitalized with severe illness, it is unlikely that many, if any, of the patients became aware of whether they were receiving the calcifediol treatment or not. The calcifediol was administered by nurses, and the treating physicians were not specifically told which patients received the treatment. The information *was* available to the treating physicians if they looked up the daily drug treatment information, but in practice it is unlikely that many of them became aware of which patients received the calcifediol.

Despite these precautions, there could have been some residual effects of imperfect blinding, such as placebo effect, experimenter bias, etc., among any patients or their treating physicians who became aware of which group they were in. However, we calculated that these residual effects would need to have been implausibly large to invalidate the conclusions of the study.

To analyze how imperfect blinding might have biased the study results toward a larger difference in ICU admissions between the treatment and control groups, we assume a “worst case” scenario, in which treated patients were less likely to be admitted to the ICU due to imperfect blinding, and control patients were more likely to be admitted to the ICU due to imperfect blinding. This fits with our intuition about the placebo effect -- if treated patients knew that they were in the treatment group, they might expect to do better than if they were unsure and if control patients knew they were *not* being treated they might expect to do worse than if they were unsure, and this expectation could influence the results; the same holds for the physicians treating them.

Suppose that some of the patients in the treatment group, or their physicians, became aware that they were in the treatment group. Let f_T_ be the fraction of treated patients in this “aware” category and let p_T_ be the probability that this awareness would “cure” the patient, i.e., the patient was not admitted to the ICU but would have been if not for this awareness. Similarly let f_C_ be the fraction of control patients or their physicians who became aware that they were *not* in the treatment group, and p_C_ be the probability that an aware patient who would not have been admitted to the ICU in a blinded study would be admitted due to this awareness. The values of p_T_ and p_C_ quantify the “unblinding effect”, namely the effect that awareness of group membership has on the need for ICU care. For any given values of f_T_, f_C_, p_T_, and p_C_, we can compute how likely it is that the results of a fully blinded study would not have been statistically significant (see Methods). To simplify, consider the case that f_T_= f_C_ and p_T_ = p_C_. Even if one third of the patients were aware which group they were in (f_T_= f_C_ = ⅓), the decreased ICU admissions that would have been observed in the treatment group in a fully blinded study would still have been expected to be statistically significant at the 95% level unless unblinding effect changed the risk of requiring ICU care by more than a factor of 7. For a given strength of the unblinding effect, we can also calculate how likely it is that randomization would distribute prognostic risk factors so unevenly that the results would no longer be statistically significant at the 95% level after adjusting for this uneven distribution *and* unblinding effect. We find that unless unblinding effect changed the risk of requiring ICU care by more than a factor of 2.1, this likelihood would be less than the usual threshold of 0.05.

We conclude that it is not plausible that the decreased ICU admissions in the treatment group were due to imperfect blinding, uneven distribution of prognostic risk factors, or a combination of the two.

### Uneven distribution of comorbidities was not significant evidence of incorrect randomization

A key assumption in our analysis has been that the randomization was truly random. It has been suggested that the significant enrichment of hypertension in the control group is evidence that it was not random [22]. The Córdoba study reported that 15 of the 26 patients in the control group (56.69%) had hypertension as compared to 11 of the 50 patients in the treatment group (24.19%) with a nominal p-value of 0.0023, meaning that random assignment of patients to the control group would only lead to an enrichment of hypertension in the control group this large 0.23% of the time. Taken by itself, this would appear to be strong evidence that the patient assignment was not random. However, there were 15 prognostic risk factors reported, and the p-value of 0.0023 was a one-tailed p-value, so there were actually 30 tests for the hypothesis, namely that one of the 15 prognostic risk factors was higher in the control group or higher in the treatment group. (Note that the hypothesis being tested here is whether the randomization was random, not whether the comorbidities could account for the difference in ICU admission; a significant difference between the two groups would be evidence that the randomization was not random irrespective of which group was enriched, which is why we need to correct for both tails.) Because no one in either group had previous kidney disease, the p-values for those two tests are not defined, leaving 28 tests with well-defined p-values. When performing multiple tests for a hypotheses, it is expected that some of them will have low nominal p-values simply due to chance; this can be corrected by applying a Bonferroni factor of 28, giving a corrected p-value of 0.0644 for the excess hypertension in the control group (see Methods). This p-value is not significant evidence that the assignment was not random at the usual 95% confidence level. This does not prove that the randomization was truly random, which is impossible in principle, it only fails to disprove it. As noted earlier, the effect of the treatment was still very large and very significant after adjusting for the excess hypertension in the control group.

### Two other randomized trials suggest early treatment is critical

The results of two additional randomized controlled trials of vitamin D formulations for COVID-19 have been reported, which help inform the interpretation of the Córdoba results.

In a North India hospital, a randomized, placebo-controlled trial for asymptomatic or mildly symptomatic SARS-CoV-2 RNA-positive vitamin D deficient (25OHD<20 ng/ml) individuals who did not require invasive ventilation or have significant comorbidities, treated 16 patients with daily 1500 mcg oral vitamin D_3_ for 7 days or until 25OHD>50 ng/ml, whereas 24 received a placebo [24]. Twelve of the treated patients achieved 25OHD>50 ng/ml by the end of the study. Ten of the 16 treated patients (62.5%) and five of the 24 controls (20.8%) became SARS-CoV-2 RNA negative after 21 days, which was the primary endpoint. The study reported a p-value of 0.018 using the two-sided Fisher exact test, but the correct p-value is 0.010 using the one-sided hypergeometric distribution because the hypothesis being tested was that treatment would decrease time to viral clearance, rather than that it would have an effect in either direction. The treatment group also showed a significant decrease in fibrinogen, but not in other inflammatory markers. The results are not entirely comparable to the Córdoba results because patients did not have severe symptoms and because treatment began immediately after diagnosis, but the study did show that high dose vitamin D_3_ can raise serum 25OHD in SARS-CoV-2-positive patients, and that early treatment with vitamin D can speed viral clearance.

In two hospitals of Sao Paulo, Brazil, a larger (n=240) double-blinded, randomized, placebo-controlled trial of vitamin D_3_ (single oral dose of 200,000 IU) for hospitalized COVID-19 patients recently reported in a medRxiv preprint [25] did not find a statistically-significant improvement in hospital length of stay, mortality, ICU admission, or need for mechanical ventilation. Serum 25OHD levels in treated patients were found to have increased from baseline by 24.0 ng/mL at discharge (average 7 days after treatment). Post hoc analysis of patients with vitamin D deficiency at baseline (n=116) showed similar results. No severe adverse reactions to vitamin D3 supplementation were reported except one patient who vomited.

The timing of when patients achieved increased 25OHD levels relative to the course of the disease might account for the disparate outcomes in the Córdoba and India studies vs. the Sao Paulo study. Patients in the Sao Paulo study were treated an average of 10.3 days after symptom onset, compared to 7.1 days for the Córdoba cohort and upon diagnosis for the India cohort. In addition to later administration, the Sao Paulo study used oral vitamin D_3_, which is known to increase serum 25OHD more slowly than calcifediol. In particular, the precise dosage and formulation of calcifediol administered in the Córdoba study, 532 mcg in soft capsules, has been found to increase serum 25OHD from less than 20 ng/ml to more than 60 ng/ml in four hours [29], whereas it took two days for 540,000 IU of vitamin D_3_(a larger dose than was used in the Sao Paulo study) to increase 25OHD from 13 ng/ml to 33.1 ng/ml [30]. These results are consistent with tests using smaller doses [31]. Thus, combining the later administration and the use of a slower-acting treatment, we infer that the effects of treatment on serum 25OHD in the Sao Paulo study were likely to have occurred an average of 12 days after symptom onset, five days later than in the Córdoba study, which might have been past the point of maximum benefit of increased 25OHD in preventing ARDS. In fact, a study of COVID-19 patients in Wuhan, China, found that among 59 patients with ARDS, median time from disease onset to ARDS was 12 days, with interquartile range 8-15 days [32].

Several other factors might have contributed to the different outcomes of these two studies, including the different treatment backgrounds. Most notably, 64.2% of treated patients in the Sao Paulo study were given corticosteroids. Steroid use is associated with low 25OHD levels [33]. Although the particular corticosteroids were not specified, dexamethasone is the corticosteroid recommended by the National Institutes for Health for COVID-19 treatment [34] and dexamethasone is known to decrease expression of *CYP2R1* [35] which encodes vitamin D 25-hydroxylase, the enzyme that is mainly responsible for converting vitamin D into 25OHD [36]. Although 25OHD levels increased on average in the treatment group by the time of discharge, the use of steroids might have delayed the increase or interfered with it in a subset of patients. Differences in patient characteristics could also have contributed; for example, 40.8% of the treated Sao Paulo patients had diabetes, as compared to only 6% of the treated Córdoba patients. Finally, although the Sao Paulo study did not find a statistically-significant effect of treatment, the 95% confidence intervals were wide, and did not rule out a decrease in relative risk of ICU admission or mortality by approximately a factor of 2.

### Will the Córdoba results generalize to other cohorts?

Showing that the decreased ICU admissions among the treated patients in the Córdoba study were associated with the calcifediol treatment is not sufficient to show that the effect will be similar in other cohorts, because the Córdoba patients might not be representative. One source of possibly relevant differences is known prognostic risk factors for the disease. To investigate this, we compared characteristics of the Córdoba cohort to those of the patients in the SOLIDARITY trial [37], a huge trial of repurposed drugs on 11,266 hospitalized COVID-19 patients in 405 hospitals in 30 countries (**Table 1**). Among the many patient characteristics reported in each study, the ones in common between the two were age, sex, lung disease, diabetes, and heart disease. Counts for the age ranges reported in the SOLIDARITY trial (<50, 50-69, and 70+) were not reported in the Córdoba study, so we estimated counts using normal distributions matching the means and standard deviations reported for the treatment and control groups. Among the characteristics reported for both studies, the Córdoba cohort had significantly fewer patients with heart disease, diabetes, and over age 70, and significantly more aged 50-69. If calcifediol treatment is less likely to help patients with heart disease or diabetes, or over age 70, or more likely to help those aged 50-69, then there could be a smaller effect in the broad SOLIDARITY cohort than in the Córdoba cohort. Furthermore, since few drugs have as large an effect size as the observed association with calcifediol in the Córdoba cohort, we would expect the effect size in other cohorts to be lower.

Another possible difference between the Córdoba cohort and other hospitalized COVID-19 patients is the treatment background. The standard of care for the Córdoba patients was hydroxychloroquine and azithromycin, which is not the current standard of care in many countries. If the effect observed in the Córdoba study was associated with a synergistic interaction between calcifediol and hydroxychloroquine or azithromycin, or if drugs in the current standard of care have an antagonistic interaction with calcifediol, then the decrease in ICU admissions might not be seen in other patients. To the extent that the standard of care has improved since mid-April 2020 when the Córdoba study was completed, the bar is higher now, so any benefit of calcifediol on top of the improved standard of care could have lower effect size.

We note that the reduced ICU admissions might not specifically relate to COVID-19. Even before COVID-19 emerged, a meta-analysis of seven randomized trials of critically ill adult patients found that vitamin D supplementation was associated with reduced mortality compared to placebo and seemed safe [38]. Of course, such effects not specific to COVID-19 would not make the treatment any less valuable for COVID-19 patients.

Our knowledge of what factors determine severity of disease for COVID-19 are incomplete, so tests in other cohorts will be required to fully assess the generalizability of the study. We would expect that the patients most likely to be helped by calcifediol treatment are those with vitamin D deficiency. Although we do not know how prevalent deficiency was in the Córdoba cohort, we do know that it is very prevalent in the groups most at risk for COVID-19, so if this is the determining factor of effectiveness then we would expect the treatment to be associated with lower ICU admissions for a substantial fraction of COVID-19 patients.

## Conclusion

In this work, we investigated several previously-proposed alternative explanations for the dramatic reduction in ICU admissions observed among hospitalized COVID-19 patients treated with calcifediol in the Córdoba study. Under the assumption of randomization, we show that chance imbalances in both reported and unreported prognostic risk factors between the treated and control groups and imperfections in the blinding process, even combined, are insufficient to explain the observed effect size differences between treated and untreated patients.

We also compared the Córdoba study parameters and results with those of two other randomized controlled trials, and propose that differences in treatment administration time and absorption rate may reconcile the observed differences in efficacy, and specifically that earlier treatment with faster absorption may provide greater benefit.

Our analysis strongly depends on the assignment to treatment and control groups being random. While it is theoretically impossible to prove randomness, we have shown that imbalances in the reported comorbidities do not provide significant evidence of non-randomness, and we are continuing to investigate whether any other factors may provide evidence of non-randomness.

Additional studies will be needed to determine how well these results generalize to other cohorts. While these issues are being worked out, the medical community should consider testing the vitamin D levels of all hospitalized COVID-19 patients, and taking remedial action for those who are deficient, to the extent that it can be done safely.

## Methods

### Proof of theorem

#### Theorem

In a randomized controlled study, let *p* be the p-value of the study results, and let *q* be the probability that randomization would distribute prognostic risk factors sufficiently unevenly between the treatment and control groups that after controlling for them the results of the study would no longer be statistically significant at the 95% level. Then 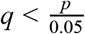.

#### Proof

Intuitively, the idea is that an extreme difference in outcomes (in this case, ICU admission) between the treated and control groups due to randomization producing an uneven distribution of prognostic risk factors is a special case of an extreme difference in outcomes due to chance, which is the p-value.

By definition, under the null hypothesis, the outcome for a patient will be the same if the patient is assigned to the treatment or control groups, so we can define an ICU patient to be a patient that will need admission to the ICU, regardless of which group the patient is assigned to. Let *U nbalancedICU* be the event that the randomization assigns ICU patients to the treatment and control groups in a way that is at least as unbalanced as what was observed in the study. By definition *p* = *Probability*(*UnbalancedICU*).

Let *M ismatchedPrognostics* be the event that a mismatch in prognostic risk factors between the treated and control groups is so large that when adjusted for it, the difference in ICU admissions between the two groups would no longer be statistically significant at the 95% level, i.e., *Probability*(*UnbalancedICU given MismatchedPrognostics*) > 0.05.

Then by the definition of conditional probability we have

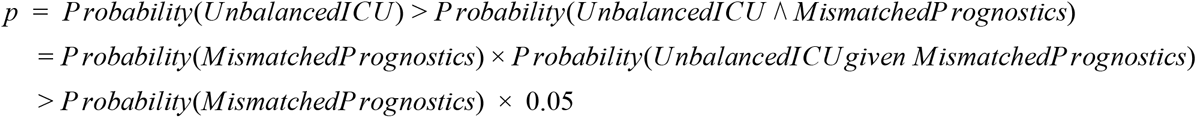

So *Probability(MismatchedP rognostics)* 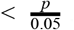.

### Effects of imperfect blinding

Define a “blinded ICU treatment patient” to be a patient in the treatment group who would have been admitted to the ICU if the study had been fully blinded (whether or not that patient was actually admitted to the ICU), and define a “blinded non-ICU control patient” to be a patient in the control group who would not have been admitted to the ICU if the study had been fully blinded (again, whether or not that patient was actually admitted to the ICU). Let *N* and *M* be the number of blinded ICU treatment patients and blinded non-ICU control patients, respectively. Let *P* be the probability that a blinded ICU treatment patient would avoid ICU admission because of imperfect blinding, and *Q* be the probability that a blinded non-ICU control patient would have been admitted to the ICU because of imperfect blinding. We have *P* = *f* _*T*_ × *p*_*T*_ and *Q* = *f* _*C*_ × *p*_*C*_ where *f* _*T*_, *f* _*C*_, *p*_*T*_, and *p*_*C*_ are the fractions of patients in each group that became aware of which group they were in and the probabilities that the awareness of an aware patient would affect that patient’s ICU admission, as defined in the main text.

The probability of obtaining the observed result that exactly one of the 50 patients in the treatment group was admitted to the ICU is the binomial coefficient *N* × *P*^*N*−1^ × (1 − *P*), and the probability of observing 13 control patients avoiding ICU admission is 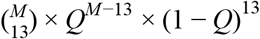.

If we assume a uniform prior on *N*, then by Bayes theorem the posterior distribution for *N* given that exactly one patient in this group required ICU admission is *C* × *N* × *P*^*N*−1^ × (1 − *P*) for some constant *C*. Since the sum of the probabilities is 1, we find that 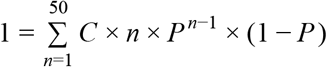, so 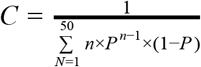 and the distribution for N is 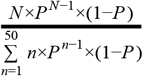. Similarly, the distribution for M is 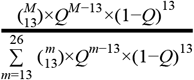. For a given *N* and *M*, the p-value of *N* out of 50 treated patients and 26 − *M* out of 26 control patients being admitted to the ICU is *h*(*N*, 50, 26, *N* + 26 − *M*) where *h* and its arguments are defined as in the R language `phyper` function, which gives the p-value for the hypergeometric distribution (the arguments are number drawn in specified category, total number in specified category, total number in other category, number drawn), so the expected p-value is 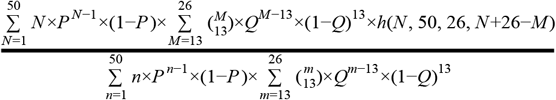.

For the case considered in the text where *P* = *Q* and 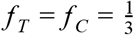, we used a standard root-finding algorithm to find that the value of *p*_*T*_ for which the p-value is 0.05 is 0.866, which corresponds to a factor of 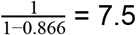 change in the risk of ICU admission for patients who were aware of their status.

Let *q*_*unblinded*_ be the probability that the randomization distributes prognostic risk factors sufficiently unevenly that, when controlling for these prognostic risk factors *and* for the effects of imperfect blinding, the result would no longer be statistically significant at the 95% level. Applying our theorem, *q*_*unblinded*_ = 0.05 is equivalent to the p-value before adjusting for this uneven distribution being 0.05 × 0.05 = 0.0025. We find that *p*_*T*_ would need to be 0.527 for the p-value to be 0.0025, which corresponds to a factor of requiring ICU admission. 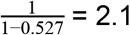 decrease in the risk of requiring ICU admission.

### Correction for multiple tests of randomness

Using a Bonferroni correction for 28 tests of randomness, we found an adjusted p-value for the enrichment of hypertension in the control group of 0.0644. An alternative would be to use the Šidák correction, which yields an adjusted p-value of 0.062. However, the Šidák correction is conservative if the tests are positively dependent and liberal if they are negatively dependent. Some of the 28 tests are positively dependent, for example age and cardiovascular disease, while others are negatively dependent, namely enrichment of any of the characteristics in the treatment group versus enrichment in the control group. Consequently it is possible that a Šidák correction would incorrectly reject the null hypothesis. In contrast, the Bonferroni correction will not incorrectly reject the null hypothesis even if the tests are dependent. In any case, the corrected values are not very different and neither falls below the usual threshold of 0.05.

### Statistical significance in Table 1

P-values in Table 1 for the difference between Córdoba and SOLIDARITY cohorts are one-sided binomial, based on percentages in the SOLIDARITY cohort. P-values in blue for the difference between calcifediol and non-calcifediol Córdoba patients were computed using the hypergeometric distribution; the ones in black are taken from Castillo et al Table 2 [21]. All p-values shown are nominal (no multiple hypothesis correction).

## Data Availability

All data is included in the paper.

## Acknowledgments

We would like to thank Michael Lev, José Manuel Quesada Gómez, Luis Manuel Entrenas-Costa, Saar Wilf, Doug Jungreis, Nick Patterson, Peter Everett, Clara Chan, Eric Meyer, and members of the online community for helpful discussions and feedback.

